# A Systematised Review of the Beighton Score Compared with Other Commonly Used Measurement Tools for Assessment and Identification of Generalised Joint Hypermobility (GJH)

**DOI:** 10.1101/2022.04.25.22274226

**Authors:** Malini Alexander

## Abstract

**Background:** 

**Objective:** A systematised review compared validity and reliability of the Beighton Score to those of other commonly used scores for identification of generalised joint hypermobility (GJH)

**Methods:** Inclusion criteria: English language, studies on humans, all types of study designs, publications in academic journals, publications from the year two thousand onwards, publications in print and theses. Exclusion criteria: studies not in English, studies measuring single joints only, studies published before the year 2000, cadaveric studies, papers with only abstracts available. An electronic literature search was undertaken of Pub Med/MEDLINE, Embase, Scopus, Cochrane Database, SPORT Discus, Pedro databases, followed by a manual search. The final review included 73 papers. The PRISMA (2021) COSMIN (2010) guidelines and CASP (2019) criteria were used to evaluate methodological quality and bias.

**Results:** The Beighton Score’s Intra-rater and inter-rater reliability ranged between ICC 0.74-0.99 and ICC 0.72-0.98 respectively. The BS has reasonable intra-rater and inter-rater reliability, however validity cannot be accurately determined as incorporation bias was identified as an issue in study methodology, not previously identified in the literature.

**Conclusion:** Paucity of data prevented accurate assessment of other scoring systems. Urgent research is required to clarify these issues and compare the BS to other tests. No source of funding was received in in undertaking this review. This review was not registered.

## 1. INTRODUCTION

Joint hypermobility can be defined as the ability of a joint to move beyond the normal range of motion. Joint hypermobility might be limited to a single joint, peripheral, axial, present in a number of joints, or widespread throughout a individual’s musculoskeletal system.

Despite prevalence of Generalised Joint Hypermobility (GJH) ranging from 2-57% in the general population, challenges with recognition using the current recommended Beighton Score exist. This leads to problems in conditions where (GJH) is the primary feature such as Hereditable Disorder of Connective Tissue including Joint Hypermobility Spectrum Disorders and Ehlers Danlos Syndrome Disorders.

Failure to recognise GJH leads to poor patient outcomes [1]. Therefore, accurate clinical assessment is of paramount importance.

This literature review compares clinimetric properties of the Beighton Score against other scoring systems used to identify GJH, including the Hospital Del Mar/Bulbena Score, The Contompasis Score, The Rotès–Quérol Score, the Upper Limb Hypermobility Assessment Tool, The Lower Limb Hypermobility Assessment Tool, the Sasche Scale and several other tools including the 5pQ questionnaire.

This paper will answer the question: How does the BS compare with other widely-used methods (including complete joint examination) for identifying GJH in patients?

Terminology in GJH literature is imprecise and ambiguous resulting in confusion in clinical practice. In this paper the terms used to describe hypermobility are taken from Castori *et al* [2]. These include the following acronyms:

- GJH – Generalised Joint Hypermobility
- JH – Joint hypermobility
- HSD – Hypermobile Spectrum Disorder
- H-EDS – Hypermobile Ehlers Danlos Syndrome
- BS – Beighton Score
- ROM – Range of motion
- HDTC – Hereditable Disorders of Connective Tissue

Interchangeable use of terms such as ligament laxity, joint hypermobility, soft tissue fragility and joint instability creates confusion for clinicians. This is potentially misleading as these terms are not anatomically equivalent [2,3]. Although closely related these clinical phenomena are influenced by a range of anatomical and physiological mechanisms. Their relationship can be conceptualised schematically in Figure 1.

**Fig. (1).**
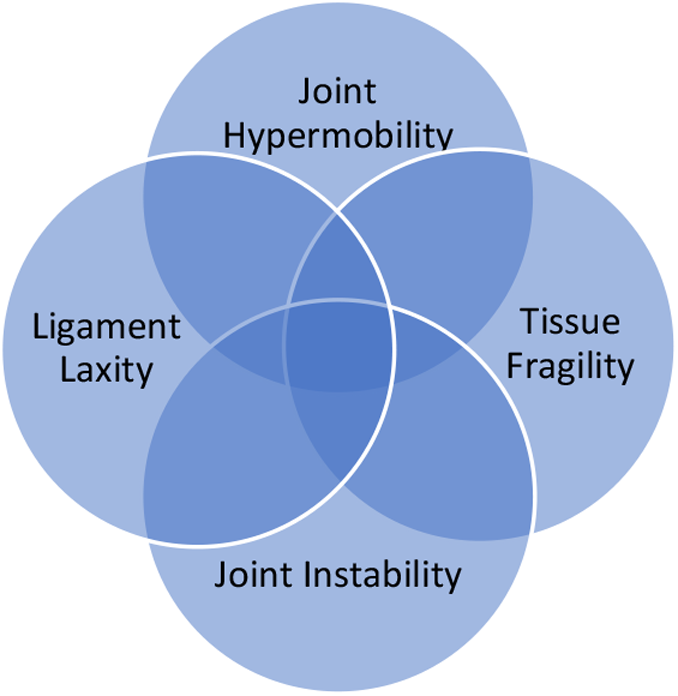
Four Closely Related, but Non-Equivalent Terms: Ligament Laxity, Joint Hypermobility, Joint Instability and Tissue Fragility.

The BS is a modification of the 1964 Carter and Wilkinson scoring system developed by Beighton and Horan in 1969 to establish prevalence of GJH in an African population [4]. It is the most commonly used method for identifying GJH.

There is no Global consensus, or gold standard cut-off score that defines joint hypermobility [5], nor is there consensus on what constitutes normal ROM therefore the very definition of what constitutes hypermobility varies in the literature as well as clinical practice, however, the Ehlers Danlos Society [6] diagnostic criteria for H-EDS endorses the paper by Castori *et al* [2], recommending a range of age-adjusted cut off scores. Use of a goniometer to assess ROM is recommended by Schlager *et al* [7], and the Ehlers Danlos Society [6].

Some researchers do not recommend the BS as the primary tool for identification of GJH due to its limitations and diagnosis should rely on a comprehensive full body assessment of joint range of motion [2,8,9]. Castori *et al* [2], and Tinkle [10] report use of any single standardized measurement tool proves challenging.

Criticisms of using the BS to establish GJH include:

- Beighton, Solomon and Soskolne [11] did not provide any evidence-based justification for the selection of joints [8]
- Only 4 joint sites are measured [8]
- Validity not adequately researched [12]
- Appropriateness for paediatric populations [13]
- Inability to capture degree of hypermobility [8]
- Developed as an epidemiological tool [8,11]
- Inclusion of ligament laxity measurement [2]
- No consensus-based cut-off values [5]
- Bias towards upper limb hypermobility, that might fail to capture lower limb hypermobility resulting in false negatives [14]
- Only assesses ROM in 2 dimensions. For some joints ROM occurs in multi-dimensions [16]
- There are no consensus values for normal ROM [15–17] and the values chosen in the BS scoring system are based on tradition, rather than evidence

## 2. MATERIALS AND METHODS

### 2.1 Databases

A single researcher undertook this review. The search strategy was conducted in accordance with recommendations of Preferred Reporting Items for Systematic Reviews and Meta-Analyses PRISMA [18]. Guidelines for systematic reviews. The author did not use any software to assist in collection of the papers and used a purely manual approach.

Databases were searched during August to November 2021 and included Pub Med/ MEDLINE, Scopus, Cochrane, Database, SPORT Discus and Pedro. The following registers were searched over the same period: Australian New Zealand Clinical Trials Registry, Clinicaltrials.gov, Centrewatch.com, ISRCTN, EU Clinical Trials Register.

A manual search of bibliographies and references from appropriate papers identified, Google and websites listed below were then manually searched to identify additional literature such as theses and publications unavailable in the databases listed above. Websites included: The Ehlers Danlos Society Website, The American College of Rheumatology, EULAR, UpToDate.com and Medscape. The University of South Wales library website was utilised to retrieve papers unavailable in research databases, or in online searches.

No additional literature was sourced by contacting authors or experts.

Studies were grouped according to experimental and quasi-experimental studies, narrative and systematic reviews and grey literature with expert opinion.

### 2.2 Inclusion Criteria

English language

Humans

Studies on adults and children

Observational studies

Cohort studies

Cross sectional studies

Randomised Control Trials

All publications in academic journals (including expert opinion)

Studies published from the year 2000 onwards Publications in print

Theses

### 2.3 Exclusion Criteria

Studies not in English, or not translated into English

Studies measuring a single joint only

Studies using the scoring system to measure a clinical presentation other than GJH, or H-EDS

Studies published before the year 2000

Grey literature

Papers for which only abstracts are available

### 2.4 Outcome measures and measures of association

Inter-rater reliability

Intra-rater reliability

Validity

Sensitivity

Specificity

Any other reported statistics relating to test properties

All forms of measurement of strength of association reported in the literature were included

### 2.5 Search Strategy and Data Extraction

Each search term was manually entered to generate a number of papers. For each search the number was added up to gain the total number for each database. Each search was independently screened and papers whose titles obviously met the eligibility criteria, or exclusion criteria were saved into desktop folders, or excluded from the search accordingly.

This process generated a high number of duplicates each time an individual database was searched. Once the search was complete, the collected papers were then screened more closely to assess eligibility and exclusion criteria and duplicates were removed and deleted.

This resulted in a final list of papers for the electronic search component. For the manual search, narrative and systematic reviews and papers from the EDS Society website were used to search bibliographies of relevant studies. These papers were searched for in Google and, or sourced from the University of South Wales Library and downloaded into folders. Searching in the University library catalogue and Google generated additional papers of interest that were screened for inclusion and exclusion criteria. Duplicates were removed.

Papers not meeting inclusion criteria, or that met exclusion criteria were deleted. This resulted in the final selection of review papers. To access each paper, if it was not available in the database where it was originally cited, then a record was made and upon completion of the review these papers were searched for in other sources such as Research Gate, Academia.edu, Deepdyve, the University of South Wales Library databases, Google Scholar and Google. If free-access full text papers could not be sourced via this process, it was assumed only the abstract was available and such studies were excluded in accordance with exclusion criteria.

This review did not cover emails, other private correspondence, or letters and errata. There are no regulatory reviews relevant to GJH as far as the author is aware.

A number of measures were reported in the literature for assessment of validity and reliability. All measures were included in the results.

GJH has been considered as a diagnosis and therefore the Critical Appraisal Skills Programme (CASP) [19] for diagnostic tests was used to assess quality of papers along with The Joanna Briggs Levels of Evidence for Diagnosis [20] to rate methodological quality of studies. Additionally, use of a goniometer, or other assessment tools was included as part of the review as it is regarded as best practice when assessing ROM [21–23].

A note was made on whether studies referenced recommended Consensus-based Standards for selection of health Measurement Instruments (COSMIN) [24] Guidelines, or any other similar standards. Systematic and narrative reviews were assessed against the updated PRISMA [18] guidelines [25]. These were summarised in a separate table included in Supplementary Material C. A high degree of heterogeneity in results was anticipated, therefore a meta-analysis was not conducted.

**Fig. (2).** Prisma flow diagram [25].

Where possible this systematic review was conducted in accordance with the PRISMA Statement: An Updated Guideline for following a Systematic Review [25] as well as the PRISMA S Statement an Extension to the Prisma Statement for Reporting Literature Searches in Systematic Reviews [26].

## 4. RESULTS

The final review consisted of 73 papers. Fifty six papers of experimental and quasi-experimental studies, 3 systematic reviews, 9 narrative reviews, and 5 grey literature and expert opinion. Selected papers from initial screening, but subsequently excluded from the final analysis were recorded and are available from the authors upon request in supplementary material B.

Results are summarised in tables. Table 3.1 provides statistical results of reliability and validity in papers that directly assessed scoring systems. Table 3.1 includes colour coded information on goniometer use, cohort and patient characteristics. Additional information includes whether assessors were blinded and whether any studies discussed their protocols with reference to the COSMIN, or other relevant guidelines.

**Table 1.**
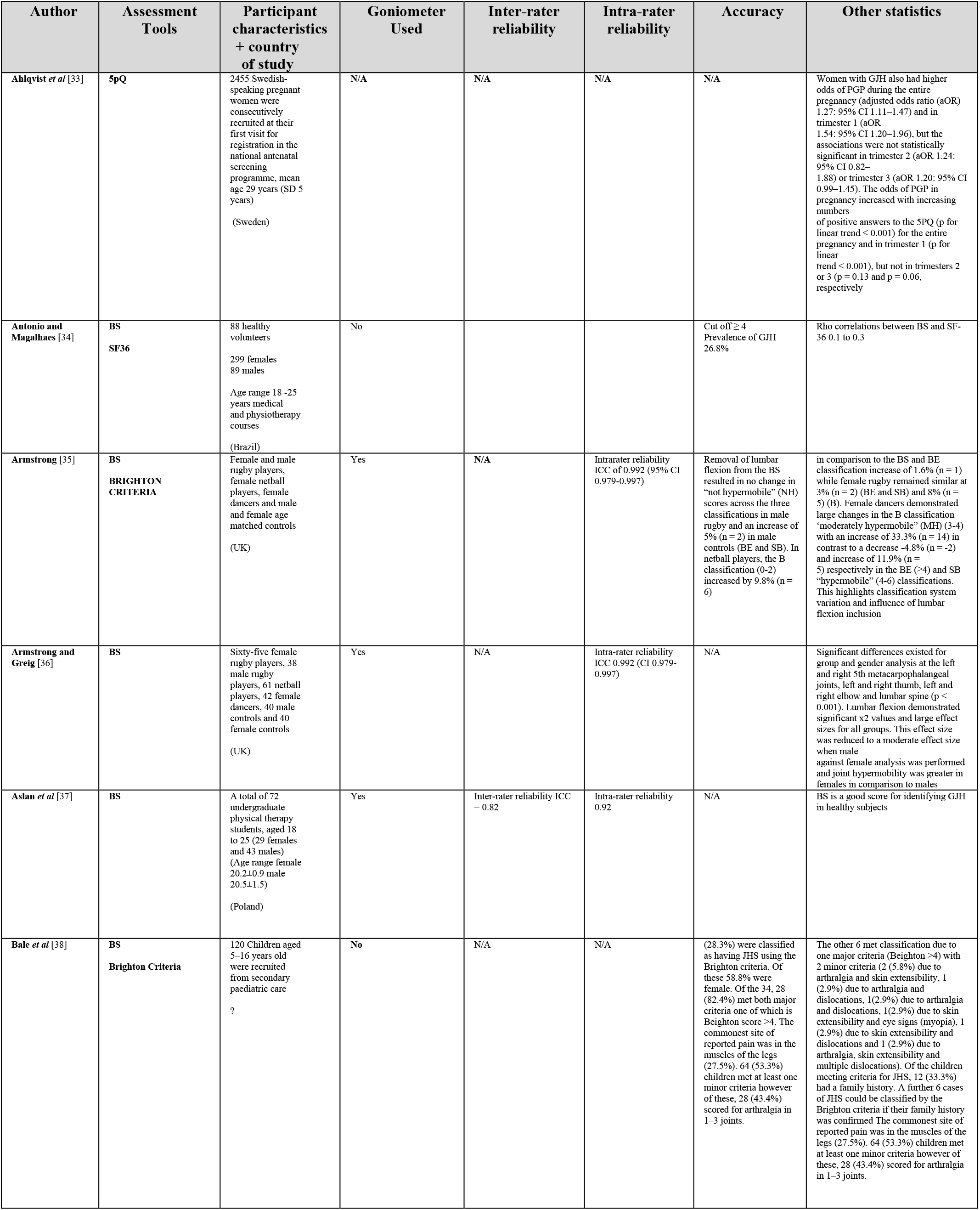
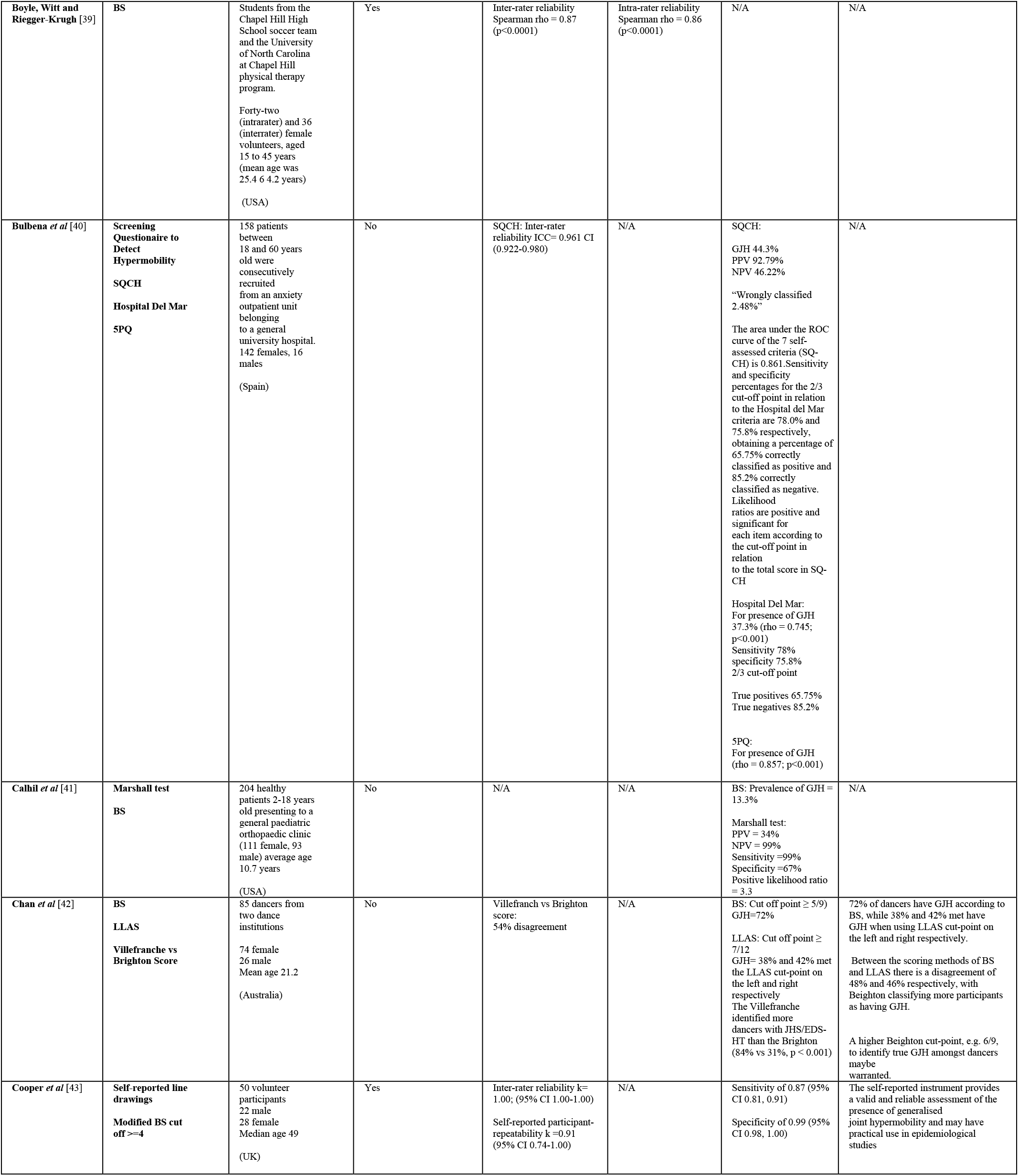
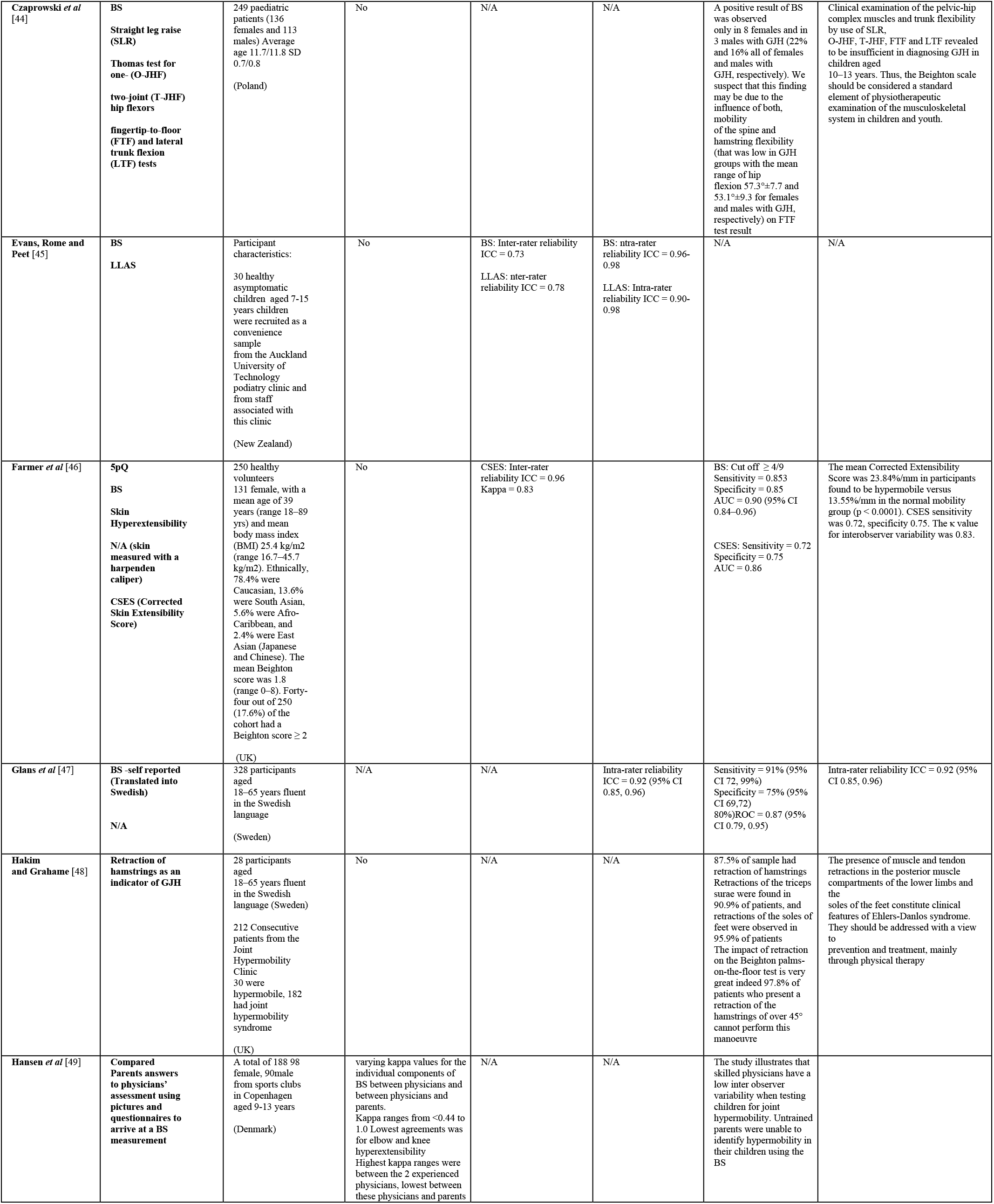
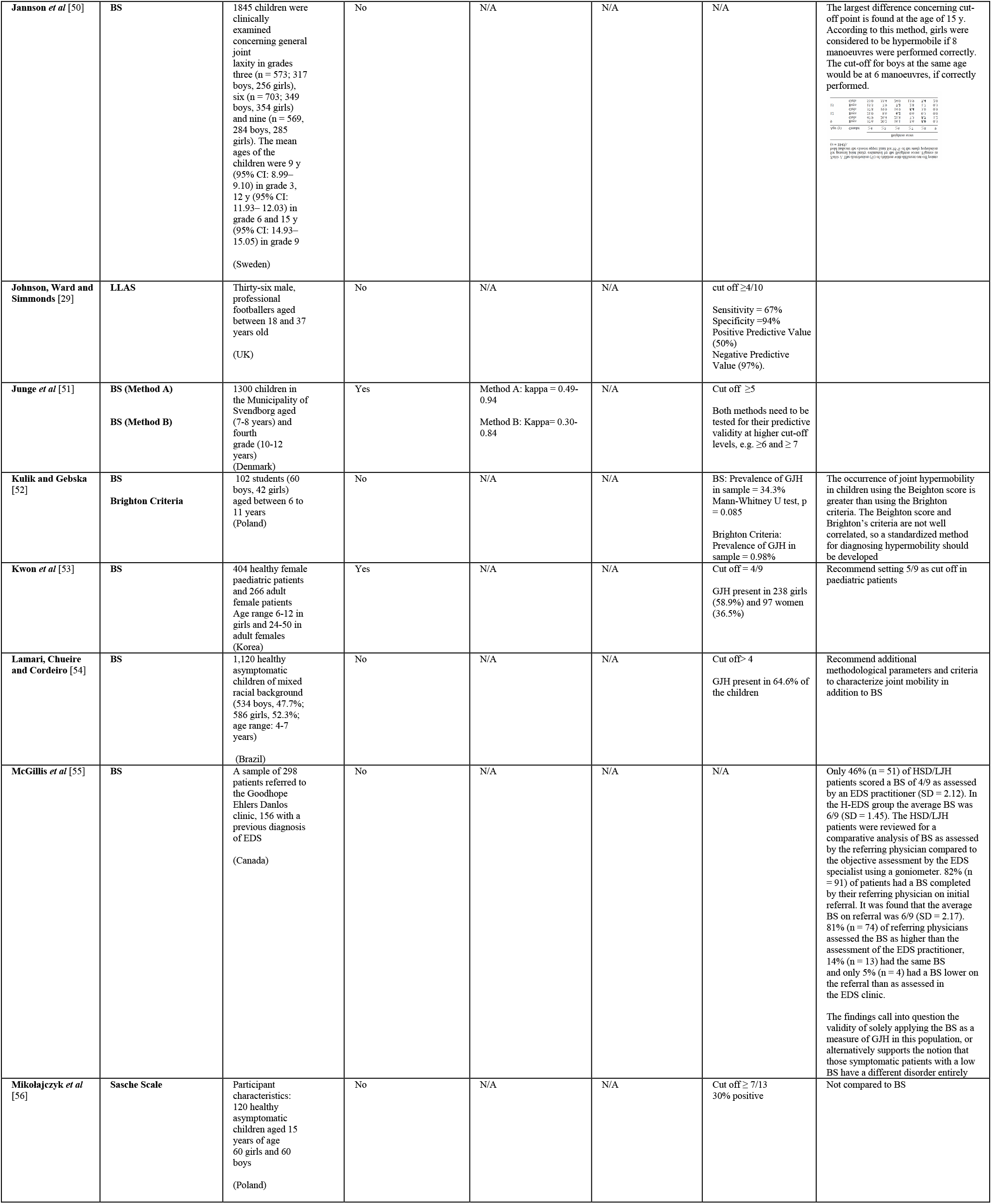
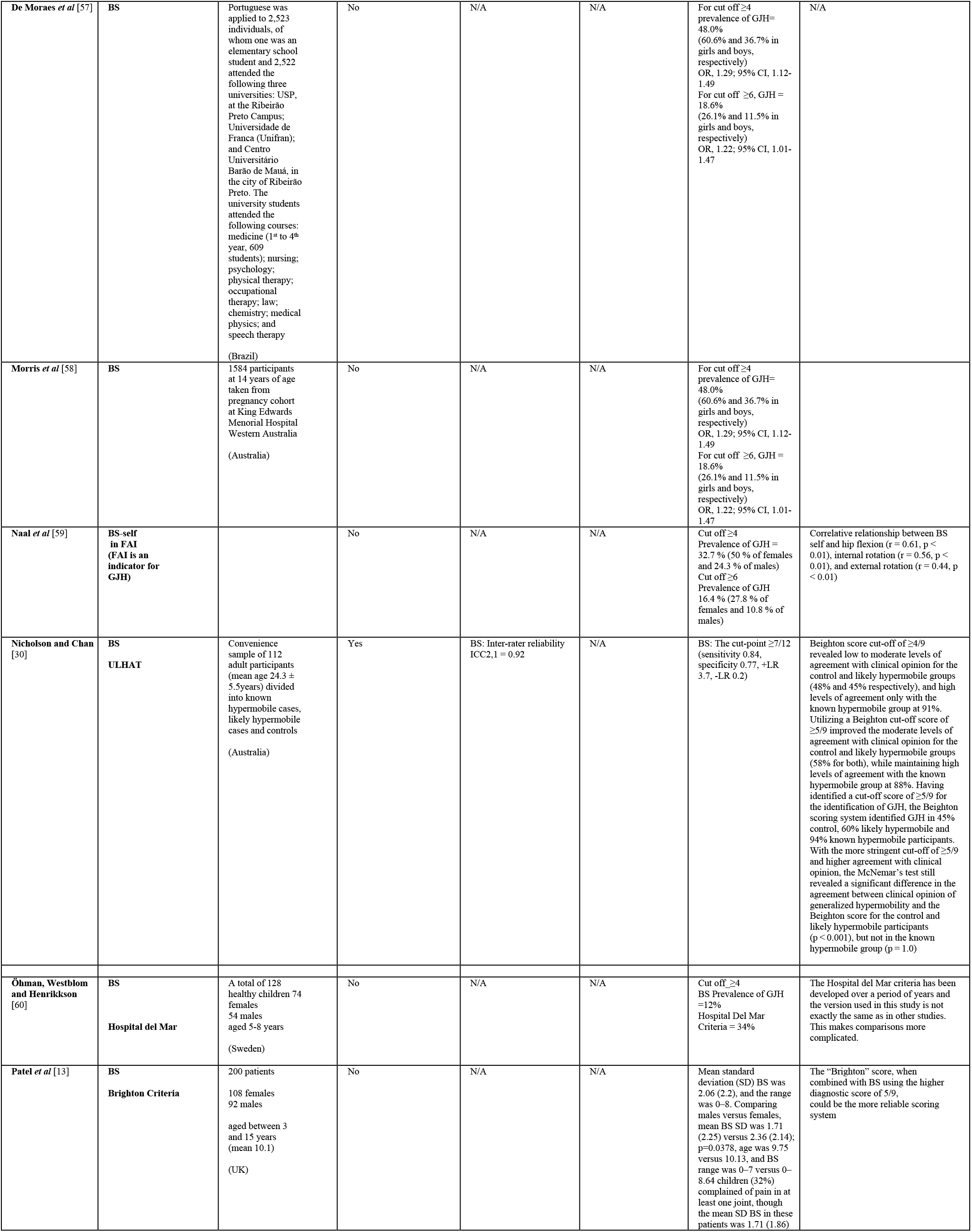
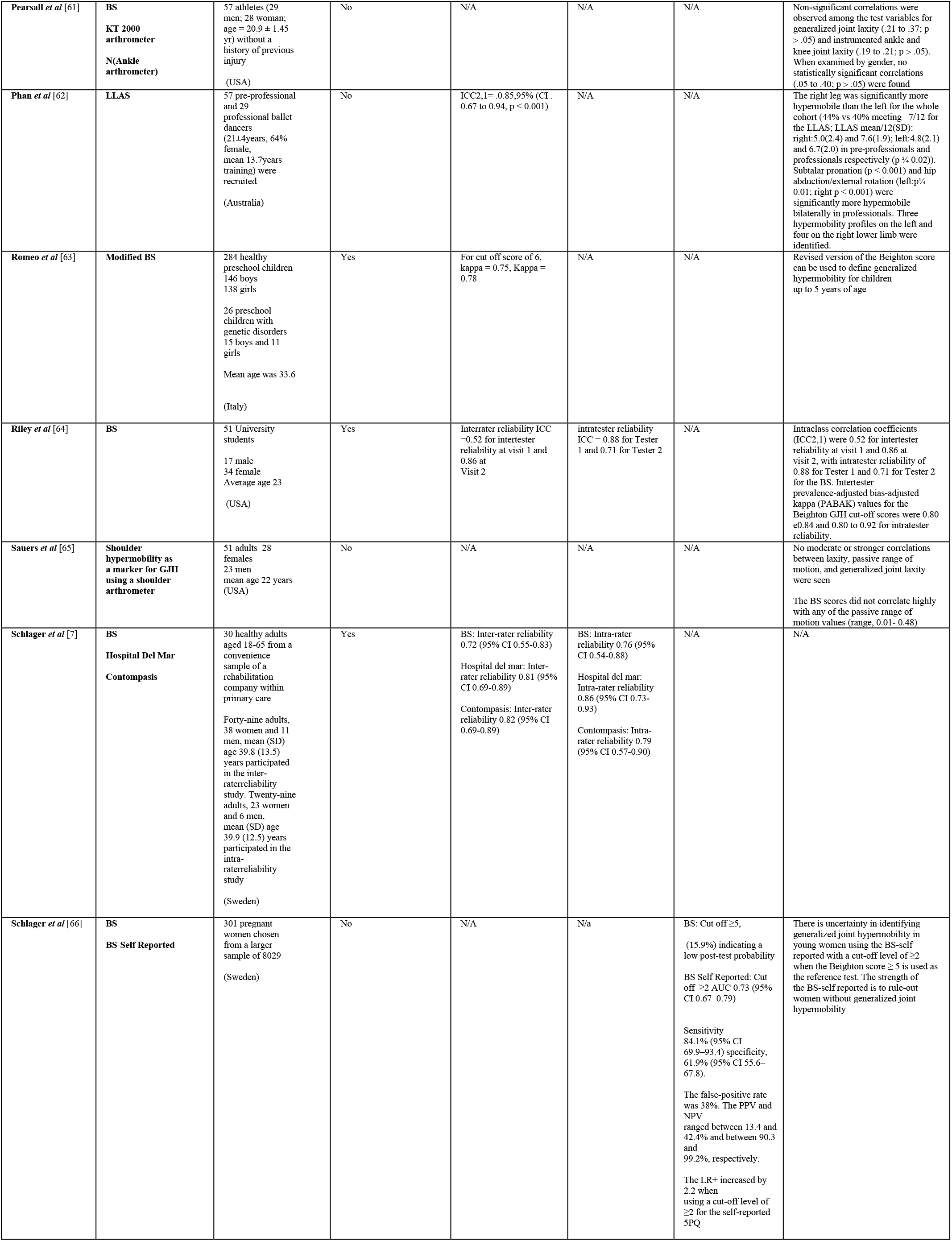
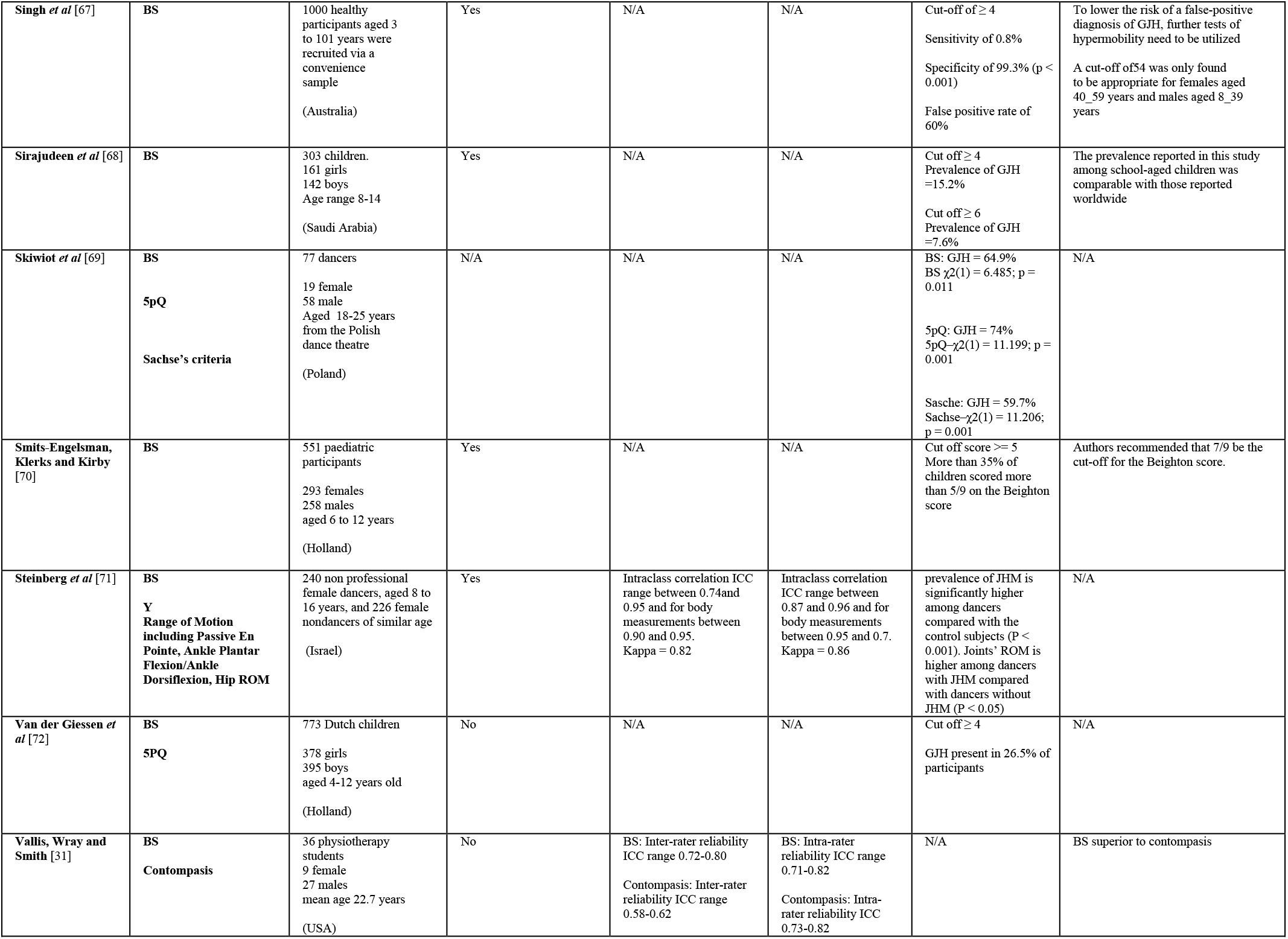
Cross Sectional Studies. JBI Level 4B.

**Table 2.**
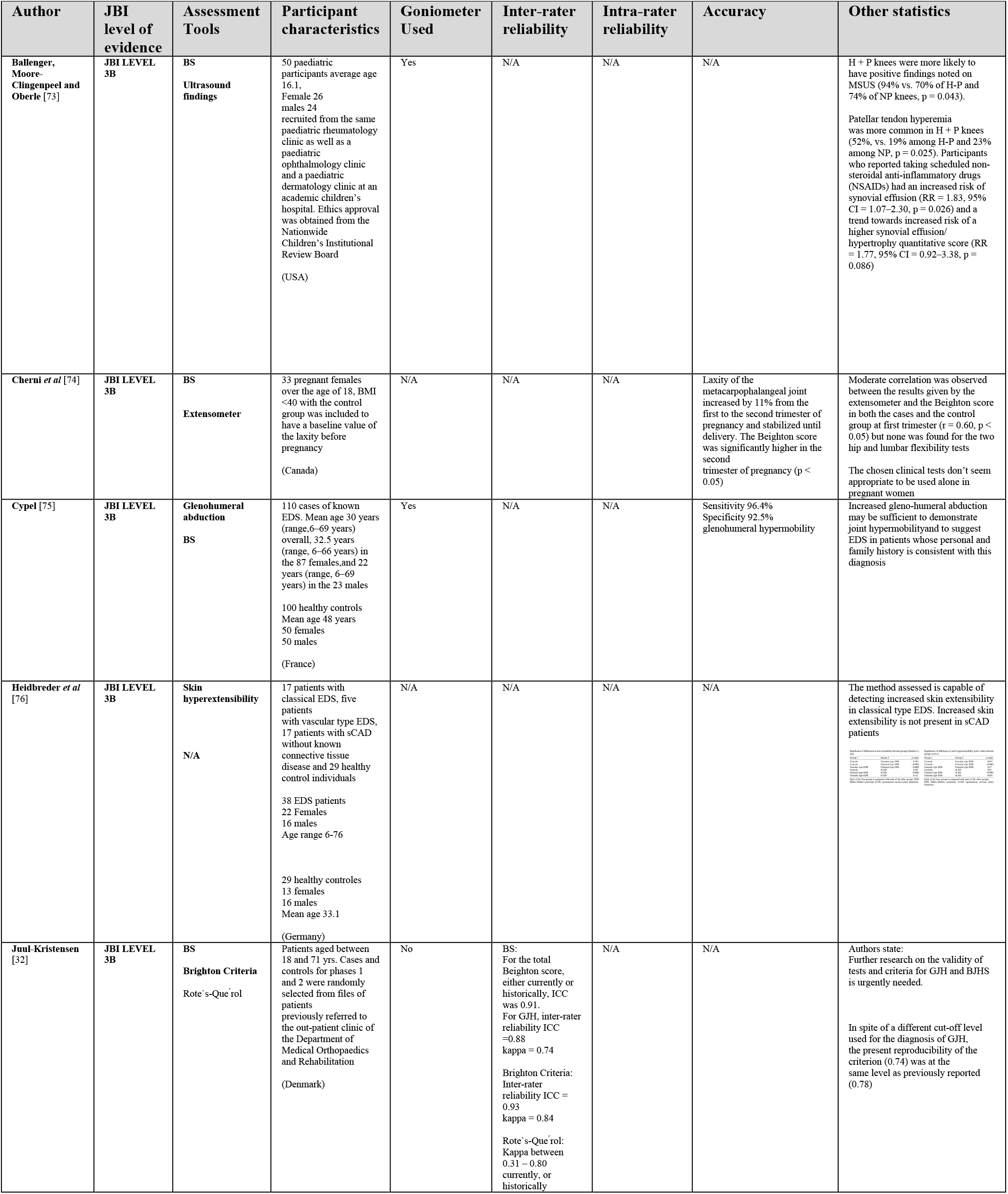
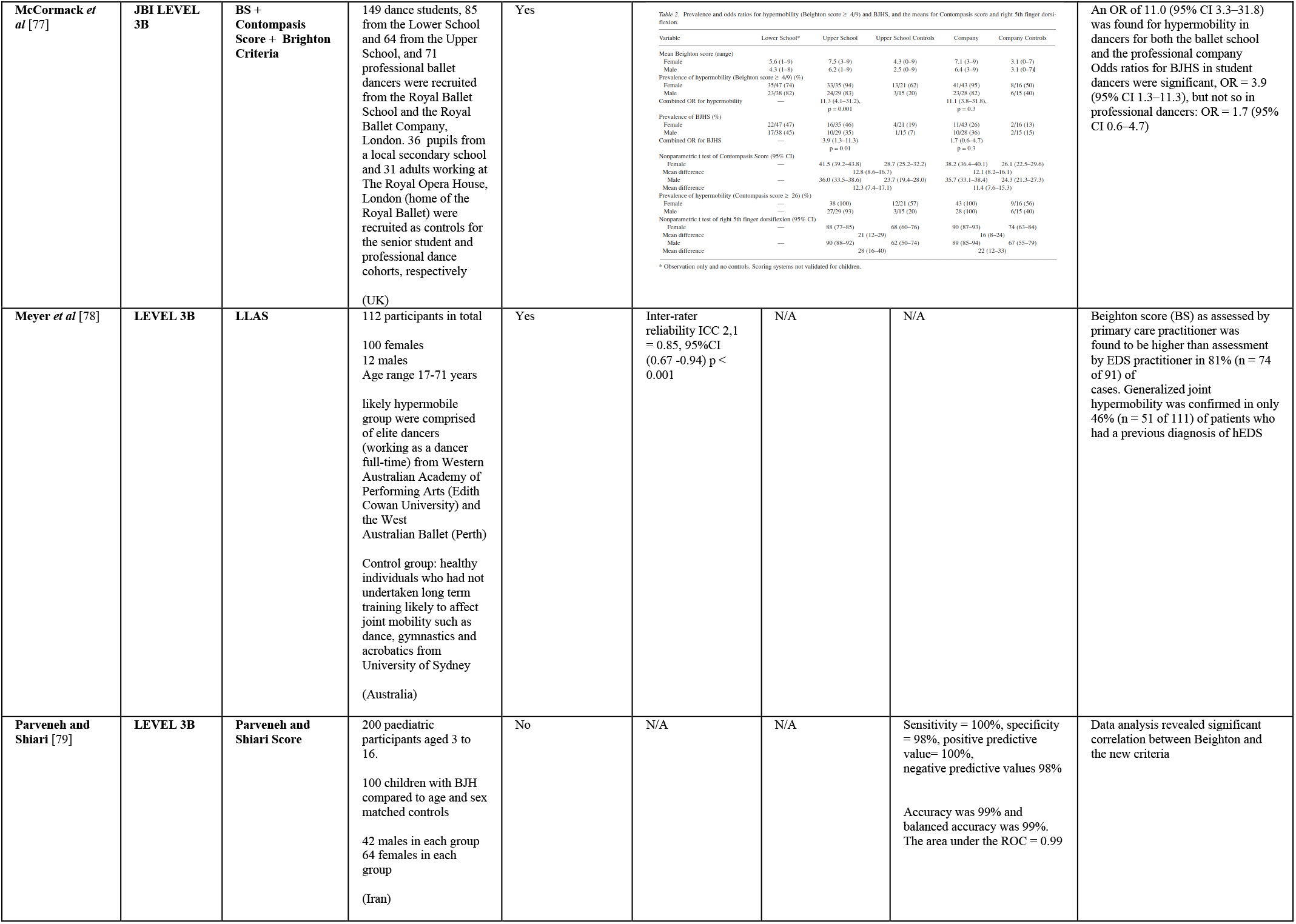
Case Control Study. JBI LEVEL 3B.

**Table 3:0.**
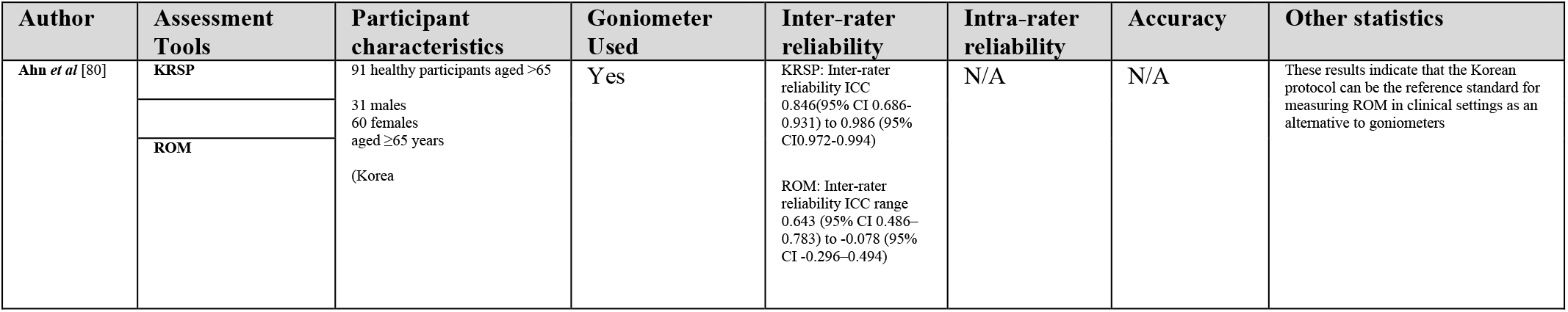
Randomised Control Trial. Level 1B.

**Table 4.**
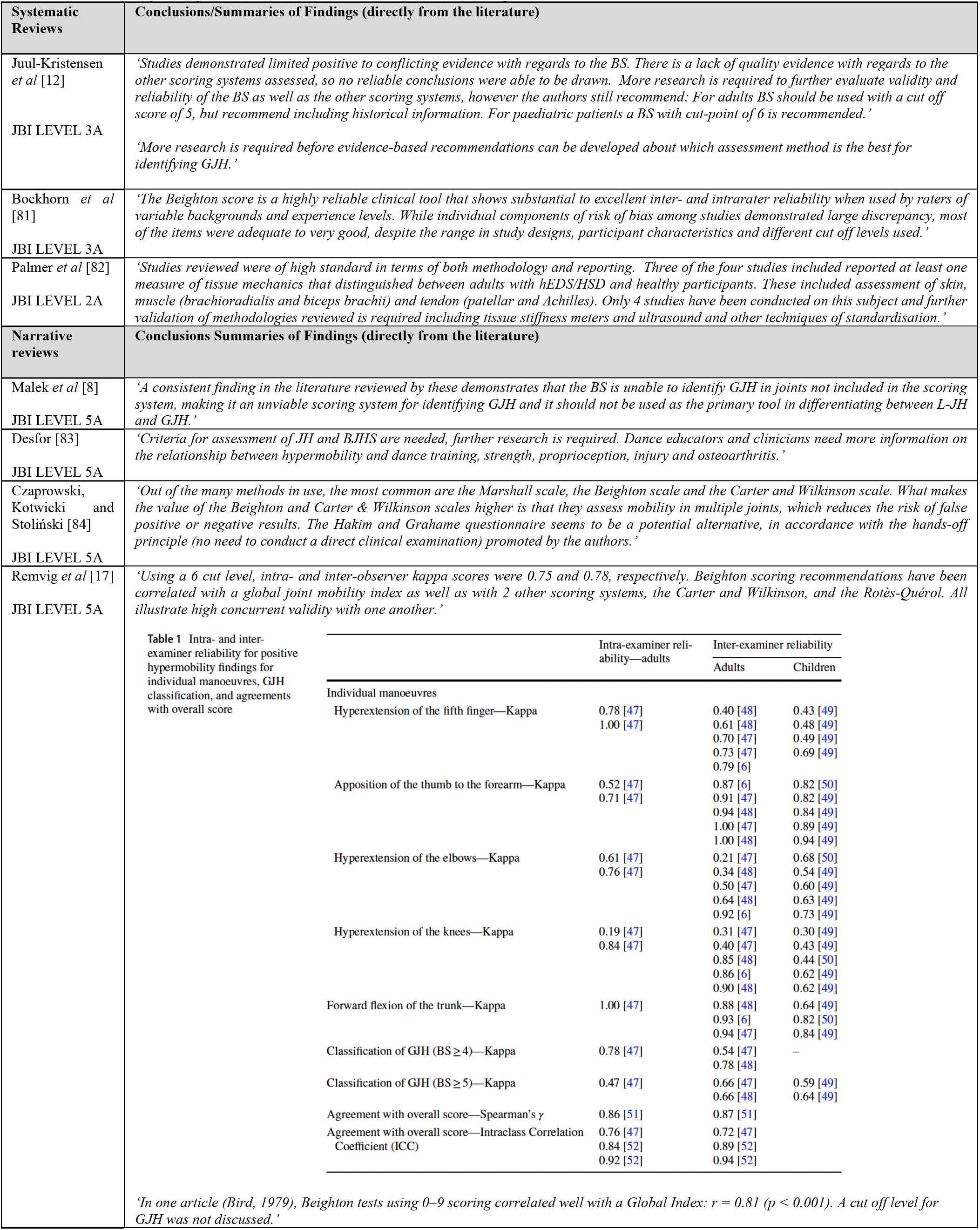
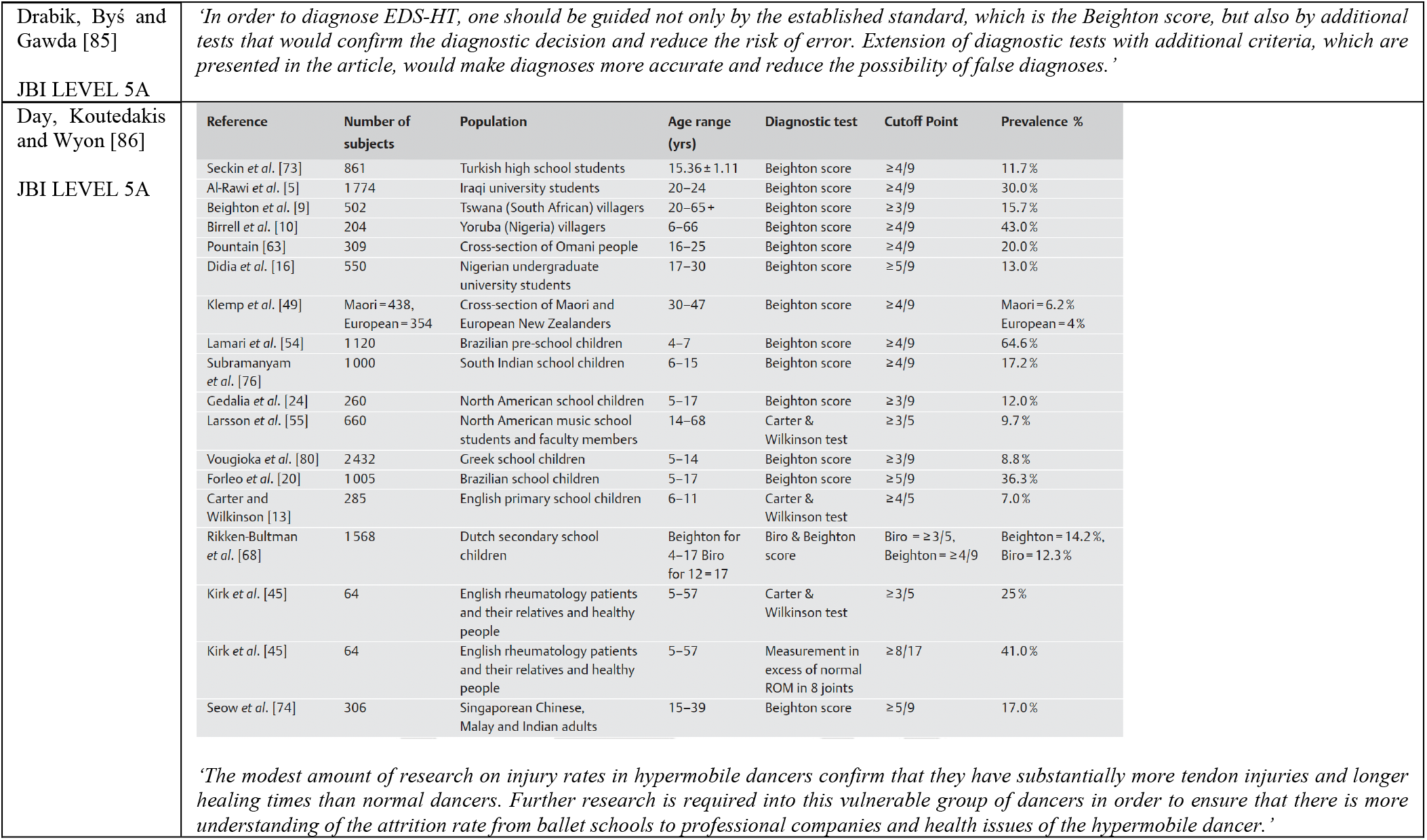
Summary of Systematic and Narrative Review Findings.

**Table 4.**
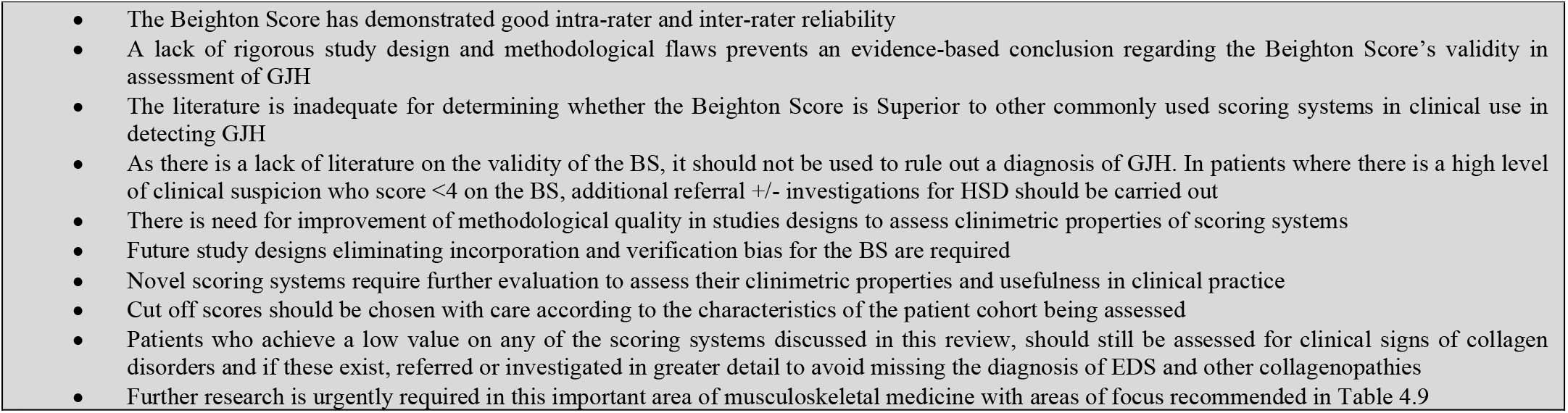
Summary of Findings.

|  |
| --- |
| • The Beighton Score has demonstrated good intra-rater and inter-rater reliability<br>• A lack of rigorous study design and methodological flaws prevents an evidence-based conclusion regarding the Beighton Score's validity in assessment of GJH<br>• The literature is inadequate for determining whether the Beighton Score is Superior to other commonly used scoring systems in clinical use in detecting GJH<br>• As there is a lack of literature on the validity of the BS, it should not be used to rule out a diagnosis of GJH. In patients where there is a high level of clinical suspicion who score <4 on the BS, additional referral +/- investigations for HSD should be carried out<br>• There is need for improvement of methodological quality in studies designs to assess clinimetric properties of scoring systems<br>• Future study designs eliminating incorporation and verification bias for the BS are required<br>• Novel scoring systems require further evaluation to assess their clinimetric properties and usefulness in clinical practice<br>• Cut off scores should be chosen with care according to the characteristics of the patient cohort being assessed<br>• Patients who achieve a low value on any of the scoring systems discussed in this review, should still be assessed for clinical signs of collagen disorders and if these exist, referred or investigated in greater detail to avoid missing the diagnosis of EDS and other collagenopathies<br>• Further research is urgently required in this important area of musculoskeletal medicine with areas of focus recommended in Table 4.9 |

**Table 5.** Grey Literature Summaries.

| Author, date, JBI Level | Type of paper | Summary of information (directly from the literature) |
| --- | --- | --- |
| Castori *et al.* [2] | Framework | *'Many EDS patients, however, still remain without a laboratory confirmation and this lack of knowledge contributes to the patients' burden. This is mostly the case for individuals affected by the two largely overlapping conditions previously termed "Ehlers–Danlos syndrome, hypermobility type (EDS-HT)" and "joint hypermobility syndrome (JHS)".'* |
| JB1 LEVEL 3B |  | *'The attribution of GJH as a feature remains partly influenced by the examiner's professional experience and recognition of the need to look at all the joints (certainly at least those in the context of the clinical presentation) and not simply those assessed in these tools'* |
| Grahame [87] | Editorial | *'The true diagnosis in a majority of patients with JHS is overlooked in up to 95% of cases.'* |
| JB1 LEVEL 5C |  | Refers to the Remvig [17] review acknowledging a lack of statistical sophistication in the majority of the papers published at the time and the need for further epidemiological studies to establish standardised joint ROM |
| Grahame [9] | Medscape article | *'Useful as an initial screen, the BS can no longer be considered the gold standard for recognizing hypermobility syndrome in clinical or epidemiological practice. It is an arbitrary all-or-none test that does not take into account the degree of laxity and in some individuals the score diminishes with advancing age, perhaps even reaching zero. Moreover, it samples only five sites, although studies have now shown that pauci-articular hypermobility is more common than polyarticular.'* |
| JB1 LEVEL 5C |  |  |
| Kacheria *et al.* [88] | Research Poster, International Association for Dental Research, AACR Annual Meeting | *'Prevalence of hypermobility using the 9 point Beighton was 12.4%, while it was 18.4% using the 4 point scale. The sensitivity was 85% and the specificity was 91%. Stratified by gender, the sensitivity and specificity were 76% and 97%, respectively, for males, and 90% and 87%, respectively, for females. The 4 point method can be accomplished in just over a minute, versus 3 minutes for the full Beighton, and the subject does not need to remove clothing or get out of a chair. Conclusions: The shortened Beighton demonstrated good sensitivity and specificity and can be done easily and quickly. It is more convenient and hence may be used as a substitute for the full Beighton scoring examination in research studies.'* |
| JB1 LEVEL 2B |  |  |
| Cincinnati Children's Hospital Medical Center Joint Hypermobility Team [89] | Clinical Practice Guidelines . Comment: Comprehensive and holistic guidelines. Incorrectly defines "joint hypermobility" as a primary diagnosis, however written prior to 2017 nosology changes recommended by Castori [2] | *'A Beighton Scale score of 5/9 or greater is used to classify JH (Scheper 2013 [1b], Junge 2013 [2b], Beighton 1998 [5a]). However, 4/9 was also frequently used to classify JH in the literature reviewed for this guideline.'* |
| JB1 LEVEL 5C |  |  |

**Table 6.** Summary Statistics: Ranges of Clinimetric Values for GJH Scores (Taken from Table 3.1 and 3.2).

| Scoring System | Intra-rater agreement range | Inter-rater agreement range | Validity Statistics range |
| --- | --- | --- | --- |
| **BS/BHJS** | ICC= range between 0.74 [71] – 0.992 [36], kappa = 0.82 [71] | ICC= 0.72 [7] - 0.96-0.98 [45]<br>Spearman rho = 0.86 (p<0.0001) [39] | The Cronbach's a for defining GJH, EDS-HT and JHS was 0.61, 0.79, and 0.44, respectively [5] |
| **Modified BS** | No data | Interrater reliability of kappa = 0.49-0.94 Method A, Kappa= 0.30-0.84 Method B [51] | No data |
| **Carter and Wilkinson Scoring System** | No data | No data | Cut off score $\geq 3 / 5$<br>GJH=25%<br>[86] |
| **Sasche Score** | No data | No data | GJH = 56.5%<br>Sachse-$\chi^2(1) = 11.206$; p = 0.001<br>[90] |
| **Brighton Criteria** | No data | Inter-rater reliability ICC = 0.93<br>kappa = 0.84 [32] | For paediatric patients:<br>GJH present in (28.3%) Of these 58.8% were female. Of the 34, 28 (82.4%) met both major criteria one of which is BS >4. [38] |
| **5pQ** | No data | ICC = 0.92 (95% CI 0.85, 0.96) [47] | Sensitivity = 70.9%<br>specificity = 77.4%<br>ROC= 0.786<br>[57]<br>Sensitivity =84%Specificity 89%<br>HSD:<br>Sensitivity=80%Specificity = 84%<br>[48]<br>presence of GJH (rho = 0.857; p<0.001)<br>[40] |
| **Line Drawings** | No data | k= 1.00; (95% CI 1.00-1.00)<br>[43] | Self-reported participant-repeatability k =0.91 (95% CI 0.74-1.00)<br>Sensitivity of 0.87 (95% CI 0.81, 0.91)<br>Specificity of 0.99 (95% CI 0.98, 1.00)<br>[43] |
| **Screening Questionnaire to Detect Joint Hypermobility** | No data | ICC= 0.961 CI (0.922-0.980) [40] | GJH 44.3%<br>PPV 92.79%<br>NPV 46.22%<br>[40] |
| **BS – self reported** | ICC = 0.92 (95% CI 0.85, 0.96) [47] | No data | Patients with FAI [59]:<br>Correlative relationship between BS-self and hip flexion (r = 0.61, p < 0.01), internal rotation (r = 0.56, p < 0.01), and external rotation (r = 0.44, p < 0.01)<br>In pregnant women [66]<br>There is uncertainty in identifying generalized joint hypermobility in young women using the BS-self reported with a cut-off level of $\geq 2$ when the Beighton score $\geq 5$ is used as the reference test. The strength of the BS-self reported is to rule-out women without generalized joint hypermobility<br>Cut off $\geq 2$ AUC 0.73 (95% CI 0.67–0.79)<br>Sensitivity 84.1% (95% CI 69.9–93.4)<br>Specificity 61.9% (95% CI 55.6–67.8)<br>The false-positive rate was 38%<br>The PPV and NPV ranged between 13.4 and 42.4% and between 90.3 and 99.2%, respectively<br>The LR+ increased by 2.2 when using a cut-off level of $\geq 2$ for the self-reported 5PQ<br>Paediatric population [47]:<br>Sensitivity = 91% (95% CI 72, 99%)<br>Specificity = 75% (95% CI 69,72)<br>80%)ROC = 0.87 (95% CI 0.79, 0.95) |
| **Marshal Test** | No data | No data | PPV = 34% |
|  |  |  | NPV = 99%<br>Sensitivity = 99%<br>Specificity = 67%<br>Positive likelihood ratio = 3.3<br>[91] |
| **KRSP** | No data | ICC 0.846- 0.986 [80] | No data |
| **Upper Limb Hypermobility Assessment Tool** | No data | ICC2,1 = 0.92 [30] | The cut-point $\geq 7/12$ (sensitivity 0.84, specificity 0.77, +LR 3.7, -LR 0.2) [30] |
| **Lower limb Hypermobility Assessment Score** | Spearman rho = 0.86 (p<0.0001) [39] | Spearman rho = 0.87 (p<0.0001) [39] | Sensitivity = 67%<br>Specificity = 94%<br>Positive Predictive Value (50%)<br>Negative Predictive Value (97%)<br>[39] |
| **Hospital Del Mar** | ICC=0.86 (95% CI 0.73-0.93) [7] | Kappa = 0.81 [92]<br>ICC = 0.81 (95% CI 0.69-0.89) [7] | For presence of GJH (rho = 0.745; p<0.001) [7] |
| **Contompasis** | ICC = 0.73-0.82 [31]<br>ICC = 0.79 (95% CI 0.57-0.90) [7] | ICC = 0.58-0.62 [31]<br>ICC = 0.82 (95% CI 0.69-0.89) [7] | No data |
| **Rotes-Querol** | No data | Kappa = 0.31 – 0.80 [32]<br>Kappa > 0.6 [17] | No data |
| **Parvaneh-Shiari Criteria** | No data | No data | Sensitivity = 100%, specificity = 98%, positive predictive value= 100%,<br>negative predictive values 98%<br>Accuracy was 99% and balanced accuracy was 99%. The area under the ROC = 0.99<br>[79] |

\*Interpretation of Agreement Scores
ICC [28] less than 0.5, between 0.5 and 0.75, between 0.75 and 0.9, and greater than 0.90 are indicative of poor, moderate, good, and excellent reliability, respectively.
Kappa [93] Minimally acceptable kappa score 0.8 or greater.
Spearman Ro [94] 1 = complete or perfect correlation, 0=no correlation
Cronbach alpha [95] >0.7 is acceptable
Chi squared If the value is greater than the significance level, then the null hypothesis is rejected

**Table 7.** Summary of Patient Cohort Information from Supplementary Material E.

| Patient Cohort | Scoring Systems Assessed | Summary |
| --- | --- | --- |
| **Pregnant Women** | BS, 5PQ, BS-Self Reported, Extensometer | BS-Self Reported good rule out test for GJH Cut off $\geq 2$ AUC 0.73 (95% CI 0.67–0.79) Sensitivity 84.1% (95% CI 69.9–93.4) specificity, 61.9% (95% CI 55.6–67.8) The false-positive rate was 38%. The PPV and NPV ranged between 13.4 and 42.4% and between 90.3 and 99.2%, respectively. The LR+ increased by 2.2 when using a cut-off level of $\geq 2$ for the self-reported 5PQ<br>Relationship between increased score in 5PQ and pelvic girdle pain during pregnancy<br>BS not good for identifying BS later in pregnancy due to inability to conduct hand to floor test |
| **Healthy Adults** | BS, Modified BS, KSRP, SQCH, 5PQ, Hospital Del Mar, Self-Reported Line Drawings, CSES, ULHAT, Shoulder Hypermobility, Hospital Del Mar, Contompasis, Rote's-Querol, Brighton Criteria, FAI | The mean Corrected Extensibility Score was 23.84%/mm in participants found to be hypermobile versus 13.55%/mm in the normal mobility group<br>ULHAT has The cut-point $\geq 7/12$ (sensitivity 0.84, specificity 0.77, +LR 3.7, -LR 0.2), Inter-rater reliability ICC2,1 = 0.92<br>No validity statistics for Rotes Querol, Kappa for interrater reliability varied between 0.31 and 0.81<br>Brighton Criteria Inter-rater reliability ICC = 0.93, kappa = 0.84<br>Patients with FAI have a prevalence of GJH = 32.7 % (50 % of females and 24.3 % of males) if using BS cut off Cut off $\geq 4$<br>BS Inter-rater reliability >0.7 in all studies except Riley *et al* [64] who report Interrater reliability ICC =0.52 for intertester reliability at visit 1 and 0.86 at Visit 2.<br>BS Intra-rater reliability >0.7 in all studies |
| **Athletes and Dancers** | BS, LLAS, Contompasis + BS + Brighton Criteria, KT arthrometer, Sasche's Criteria, Villefranche, | BS Inter-rater reliability ICC>0.7 in all studies, Intra-rater reliability>0.87, but only 2 studies looked at this statistic<br>GJH>60% using BS cut off $\geq 4$ in all studies assessing prevalence<br>LLAS demonstrated GJH= 38% with cut off score $\geq 7/12$<br>The Villefranche Criteria identified more dancers with JHS/EDS-HT than the Brighton (84% vs 31%, p < 0.001) |
| **Paediatrics** | BS, Modified BS, Hospital Del Mar, Marshall Test, LLAS, BS Self Reported in Swedish, Sasche Scale, 5PQ, Brighton Criteria, Parvaneer and Shiari Score | Hospital Del Mar GJH ranges between 38% and 70%, Inter-rater reliability Kappa = 0.81 PPV = 34% NPV = 99% Sensitivity = 99% Specificity = 67% Positive likelihood ratio = 3.3<br>LLAS Inter-rater reliability ICC = 0.78 , Intra-rater reliability ICC = 0.90-0.98<br>BS self reported sensitivity = 91% (95% CI 72, 99%) Specificity = 75% (95% CI 69,72) 80%) ROC = 0.87 (95% CI 0.79, 0.95)<br>Sasche Scale Cut off $\geq 7/13$ (30% positive)<br>BS cut off Cut off $\geq 4$ Prevalence of GJH ranged from 12% to 64.6%<br>BS Cut off $\geq 6$ GJH ranged from 7.6% to 18.6%<br>Parvaneh and Shiari Score Sensitivity = 100%, specificity = 98%, positive predictive value= 100%, negative predictive values 98% Accuracy was 99% and balanced accuracy was 99%. The area under the ROC = 0.99<br>Brighton Score Prevalence of GJH in sample = 0.98% (in healthy sample) vs 28.3% in secondary paediatric care sample<br>BS Self-Reported (in Swedish) Intra-rater reliability ICC = 0.92 (95% CI 0.85, 0.96) , sensitivity = 91% (95% CI 72, 99%) Specificity = 75% (95% CI 69,72-80%) ROC = 0.87 (95% CI 0.79, 0.95)<br>Marshall Test PPV = 34% NPV = 99% Sensitivity = 99% Specificity = 67% Positive likelihood ratio = 3.3 |
| EDS/HSD Patients | Glenohumeral abduction + BS, Retraction of tendons, BS | 87.5% of sample had retraction of hamstrings<br>Retractions of the triceps surae were found in 90.9% of patients, and retractions of the soles of feet were observed in 95.9% of patients<br>The impact of retraction on the Beighton palms-on-the-floor test is very great indeed 97.8% of patients who present a retraction of the hamstrings of over 45° cannot perform this manoeuvre, which will affect the BS and potentially identification of the condition in this cohort<br>Skin hypermobility statistically significant for classical EDS patients<br>Only 46% HSD/LJH patients scored a BS of 4/9 as assessed by an EDS practitioner (SD = 2.12). In the H-EDS group the average BS was 6/9 (SD = 1.45). |

The Joanna Briggs Level of Evidence [20] for each study is reported. The majority were rated as a Level 2b-3b. Many studies did not report whether consecutive patients were assessed. If not reported it was assumed they were not consecutive patient studies.

If a paper did not explicitly state goniometer use, it was assumed none was. If a paper did not explicitly state whether blinding of researchers, or participants was undertaken, it was assumed no blinding occurred. Blinding in the context of these research scores is with regards to participants knowing which scoring system is being used, or whether examiners were aware of each other’s scoring points, or what patients had been scored as previously.

Table 3.2 is a summary of the narrative and systematic review findings. Table 3.3 is table of expert opinion, or grey literature sources included in the study.

The tables where methodological quality was assessed according to the CASP [19] diagnostic tests and the PRISMA [18] checklist for reporting Systematic Reviews are included in Supplementary Material C and D respectively.

## 5. DISCUSSION

Systematic reviews, mainly included cross sectional studies, some included reviews of case control studies and were therefore given a rating of 3A instead of 2A. Narrative reviews rated 5A as per recommendations from the University of Canberra [27] suggesting narrative reviews are to be considered as expert opinion.

The PRISMA (2020) [25] checklist was used to evaluate both systematic reviews and narrative literature reviews to demonstrate any methodological flaws and group the reviews together.

Certain studies were designed to be association studies, rather than studies focusing specifically on clinimetric statistics and these were excluded from the review.

A statistical synthesis of results was not conducted due to heterogeneity.

Interpretation of reliability statistics can be problematic and non-straightforward [28]. Wide variation is reported for clinimetric statistics with no standardised reporting making direct comparisons difficult. This includes interpretation of effect sizes. Few papers addressed strength of association.

There was inconsistency of reporting inter-rater reliability, but not intra-rater reliability. Only some papers assessed validity and no papers assessed responsiveness.

Compared to the BS there is sparce literature on clinimetric properties of alternative scoring systems.

Intra-rater reliability scores for the BS range from ICC 0.74-0.99, K = 0.82. Inter-rater reliability score ranged from ICC 0.72-0.98, spearman rho = 0.86. For the other scoring systems assessed the score ranges are summarised in table 3.

Johnson, Ward and Simmonds [29] found a strong correlation between LLAS and BS scores (rho = 0.732; p < 0.001). The ULHAS had an intra-rater reliability ICC of 0.92 [30]. No data is available on inter-rater reliability, or validity and so a direct comparison between ULHAS and the BS cannot be made.

Vallis, Wray and Smith [31] found both inter-rater reliability and intra-rater reliability of Contompasis inferior to those of BS Inter-rater reliability ICC range 0.58-0.62 and intra-rater reliability ICC 0.73-0.82 versus Inter-rater reliability ICC range 0.72-0.80, intra-rater reliability ICC range 0.71-0.82, However Schlager *et al* [7] found both inter-rater reliability and intra-rater reliability of BS inferior to that of Contompasis Inter-rater reliability 0.82 (95% CI 0.69-0.89) and Intra-rater reliability 0.79 (95% CI 0.57-0.90) vs Inter-rater reliability 0.72 (95% CI 0.55-0.83) and Intra-rater reliability 0.76 (95% CI 0.54-0.88), respectively. Both these studies were performed in healthy adult populations, with Vallis Wray and Smith [31] selecting the sample from physiotherapy students at a university.

There was only a single paper for reliability statistics for the Rotes-Querol Score by Juul-Kristensen *et al* [32] reporting a Kappa range of 0.31-0.80. This range is lower than the Parvaneh and Shiari [79] report a perfect sensitivity of 100% for their score which is essentially a modified BS. There are no reliability statistics available for this scoring system.

Bulbena *et al* [40] found the self-reported questionnaire positively correlates with both the Hospital del Mar criteria (rho = 0. 745; p<0.001) and the self-reporting questionnaire of Hakim and Grahame [48] (rho = 0.857; p<0.001).

Schlager *et al* [66] found the Hospital del Mar Criteria superior to the BS with regards to both inter-rater and intrarater reliability statistics (Inter-rater reliability 0.81 (95% CI 0.69-0.89) and Intra-rater reliability 0.86 (95% CI 0.73-0.93) versus Intra-rater reliability 0.76 (95% CI 0.54-0.88) and Intra-rater reliability 0.76 (95% CI 0.54-0.88) respectively. Using a cut off score of ≥4 Öhman, Westblom and Henrikkson [60] found the Hospital Del Mar Criteria superior in its ability to detect GJH at 34% versus 12% for the BS.

No reliability data was available for the Sasche Scale. Skwiot *et al* [90] reported an ability to detect GJH in 56.5% of participants with a χ2(1) = 11.206; p = 0.001 (no cut off score mentioned), however Mikołajczyk *et al* [56] reported the Sasche Scale’s ability to detect GJH as significantly lower at 30% using a cut off score of ≥ 7/13. This large variance is likely to be due to differences in patient cohorts.

The Rotes-Querol scoring system is the only test that provides non dichotomous scores for hypermobility.

Calhil *et al* [41] found the Marshall test a good rule out test with specificity of 99%, but low sensitivity of 67%.

The BS is brought into question for identifying GJH in the H-EDS patient group by McGillis *et al* [55]. This is the principle method for screening H-EDS when combined with the 5pQ as per the EDS (2017) [6] diagnostic criteria. Joint hypermobility is regarded as the hallmark feature of H-EDS and other EDS subtypes, however authors McGillis *et al* [55], Hamonet and Brock [96] and Malek *et al* [8] report many patients with EDS do not always score highly on the BS.

A reason not identified in current literature is the fact EDS is a disorder of collagen fragility (hence it’s systemic nature), however hypermobility has traditionally been utilised as a proxy measure of tissue fragility. As discussed in the introduction, joint hypermobility, whilst related to tissue fragility, is not one in the same phenomenon.

This raises a question regarding nosology of HSD and whether this is truly a condition of hypermobility, or whether it is a disorder of tissue fragility and whether “Connective Tissue Fragility Spectrum Disorders”, or similar term is worth considering as alternative nomenclature. It also raises the question in patients who present with clear features of systemic tissue fragility without joint hypermobility should be considered part of this spectrum. Currently such patients do not fall into either the HSD, or EDS nomenclature, but clearly present with connective tissue pathology.

An additional question arises about whether it is possible for some patients to have tissue fragility in the absence of GJH. Currently it is not possible to identify such patients who might be currently misclassified as HSD, or not classified at all.

Degrees of consensus around reliability and validity statistics were reported in systematic and narrative reviews with conflicting conclusions about the usefulness of the BS in detecting GJH.

The majority of reviews including Remvig *et al* [17], Juul-Kristensen *et al* [12], Malek *et al* [8] and Bockhorn *et al* [81] included a high number of association studies. The author determined many of these are not strictly related to clinimetric properties of the scoring systems analysed and therefore should not be included in a review primarily focused on reliability and validity statistics.

Remvig *et al* [17] conclude all scores are comparable, however report the Rotes-Querol has a lower Kappa value compared with other scoring systems assessed.

Malek *et al* [8] conclude use of the BS as a clinical diagnostic tool, particularly within the 2017 International Classification of EDS for the diagnosis of hypermobile EDS (hEDS), remains controversial.

Juul-Kristensen *et al* [12] conclude evidence supports the BS as a reliable clinical tool, however are concerned with a lack of research relating to validity. There is insufficient data to draw conclusions about the other scoring methods assessed in their review.

Drabik, Byś and Gawda [85] report GJH and H-EDS might be more accurately identified when combining the BS with other assessments.

Day, Koutedakis, and Wyon [86] reviewed hypermobility in dance including papers relating to dancers and non-dancers with a difference in detection rates between the Carter and Wilkinson method vs the Beighton Score, however these studies included different types of participants it is difficult to ascertain whether detection rates differ as a result of participant characteristics, or difference in the ability of scores to detect GJH.

Bockhorn *et al* [81] report the BS is excellent in terms of reliability. This review states publication bias and the lack of standardised reporting with use of either composite scores, individual measurements, or variable cut off scores could lead researchers to be influenced to choose the value with the highest level of agreement.

Lack of research to establish validity of the BS raised by Remvig *et al* [17], Malek *et al* [8] and Juul-Kristensen *et al* [12] should be addressed as a priority area in future research.

Most reviews agreed there is lack of high-quality studies, with the exception of the paper by Palmer *et al* [82] a different patient cohort from most papers (known EDS/HSD), hence only four high quality studies were included in their review.

Few papers are high quality. Methodological quality issues include failure to use, or report use of gold standard goniometry devices for assessment of ROM, non-representative patient samples, convenience sampling, non-blinding of participants and assessors, lack of appropriate statistical analysis and, or reporting and failure to compare diagnostic studies to the recommended best practice guidelines. Small sample size is potentially an issue in the studies. Many papers used sample sizes of less than 100, the average sample size ranged from 100-300. 6 studies included a sample size greater than 1000. Some studies did not discuss what cut-off score was used, whilst others did not explain the rationale of cut-off score choice.

In establishing diagnoses, clinicians should rely on evidence-based diagnostic methodology that is standardised according to the COSMIN 2010 [24] criteria, satisfying 3 domains of validity, reliability and responsiveness (the test’s ability to detect change over time) [97]. None of the studies reviewed looks at all three of these domains.

Few papers referenced appropriate reporting, or test design standards such as STARD [98] or QUADAS-2 [99]. Some studies were conducted prior to authorship of the COSMIN [24] guidelines, however there is a general lack of referencing to other appropriate standards. Riley *et al* [64] reference the Guidelines for Reporting Reliability and Agreement Studies (GRRAS).

As no standardised reporting of clinimetric statistics exists, comparison is challenging.

Only one randomised control trial (RCT) was identified; Ahn *et al* [80], however this only partially adhered to standards for conducting a RCT.

Specific patient characteristics possibly impacted results of the BS ability to detect GJH. For example, in athlete and dance cohorts. The results and analysis of a study by Riley *et al* [64] noted prevalence of GJH within a sample has an impact on the kappa values of this tool. As reported by Lijmer *et al* [100] diagnostic studies with methodological flaws possibly over-estimate the accuracy of a test and falsely increase the pre-test probability.

A challenge in assessment of validity and reliability studies with regards to the BS, is the fact that most prevalence studies use the BS score itself to establish the presence of GJH. This introduces an inherent level of bias as the scoring method is essentially tested against itself.

This bias is likely to overestimate the sensitivity and specificity values, because these have been based on prevalence calculated using the same test. This is a source of major bias and flaw in study methodology in validity assessment of the BS. This is discussed as a source of bias in validity studies of diagnostic test accuracy by Worster and Carpenter [101] and Kea, Hall and Wang [102]. Incorporation bias would also impact the cut-off values for tests in different cohorts and is an area that requires serious review.

Paucity of literature exists on whether a combination of tests would improve the validity of scoring systems. It is possible the BS in conjunction with other methods yields a higher sensitivity compared with a single scoring system [85,30,54].

### 5.1 Heterogeneity of Results

High heterogeneity as well as conflicting conclusions drawn by researchers is present.

The complexity of subject material and lack of standardisation of procedures and protocols in clinical practice and current literature are some reasons for heterogeneity.

Other causes of heterogeneity include:

- High variability in study design methodology (case-control, vs cohort study, vs trial vs review)
- Comparison of different range of scores (not all studies compared the same scores, not all studies compared the scores with the gold standard, or included a full clinical assessment)
- Use of different cut off score (most papers use between 4-5 in adult populations and 6-7 in paediatric papers)
- Failure to use goniometer
- Lack of standardised reporting methods
- Lack of standardised statistical parameters for validity and reliability (kappa vs cronbach alpha, vs ICC)
- No standardised protocol for performing BS
- Wide differences in participant characteristics (children, vs pregnant women, vs adults, vs dancers, vs elite athletes)
- Wide differences in study size

### 5.2 Limitations and Bias in this Review

The single reviewer created possible selection bias and lack of use of systematic review software are the two most important limitations of this review. Other limitations include the search strategy, that did not use meta search terms and did not use the term “psychometric properties” when conducting the search.

The variety of non-standardised terminology potentially poses an issue, however no critical studies have been missed from the review that would alter findings due to a sufficiently broad search criteria and thorough manual search.

Other sources of bias and limitations of this paper include:

- Manual download of papers creates possible reproducibility issues
- Methodological quality of papers used in the review
- Recommended software for conducting a systematic review was not used
- Limitations of inclusion and exclusion criteria
- Broad inclusion criteria
- Variability of reported results
- Limited review of grey literature
- Limited review clinical practice guidelines
- Inclusion of some low-quality papers, grey literature and narrative reports
- English only publications
- Limited to 4 databases
- Only free papers were accessed

Areas for future research should focus on the following areas:

- Standardisation of protocols
- Evaluation between tip of thumb vs more of thumb approximation to forearm
- Consensus on cut off scores
- Lack of individual studies assessing BS specifically for COSMIN [24] criteria for validation
- Explore alternative scores for patients with tight hamstrings -> hand to floor measurement
- Explore alternative scores for patients with ligament laxity as opposed to joint hypermobility
- Whether combining BS with other scoring systems provides increased reliability and validity of findings
- The need to establish a consensus-based gold standard test (or combination of tests) for recognition of GJH
- Investigation of a standardised “global index of joint hypermobility”
- Appropriate study designs to further investigate and compare validity, reproducibility and reliability properties of all widely used scoring systems including questionnaires as these potentially have numerous benefits in a clinical context for the assessment of GJH, including reduced time spent assessing a patient without an examination and the ability to be conducted prior to appointments
- More studies on the Rotes Querol scoring system that measures degrees of hypermobility, rather than the currently dichotomous scoring systems
- A study to determine appropriate statistically derived cut-off scores for the general population vs subgroups such as professional dancers and athletes where a high pre-test probability is likely to affect the scoring systems utilised and affect interpretation of validity and reliability
- A study designed to assess joint ROM in patients with comorbid arthritis with adjusted scoring systems
- Consensus on ROM measurements for hypermobility
- Research into combined assessment methods for establishing a diagnosis of GJH vs single scoring systems
- Systematic review of instruments, software and devices for measurement of joint ROM
- Systematic review of novel associations between various clinical signs and GJH Standardised reporting methods in accordance with the many established guidelines on assessing diagnostic tests, such as COSMIN [24].

## CONCLUSION

The Beighton Score is the most commonly used score for identify GJH. This review compared reliability and validity statistics of BS to other scores for identifying BS.

The findings of this systematised review support those of previous similar reviews conducted by Juul-Kristensen *et al* [12], Bockhorn *et al* [81], and Malek *et al* [8] and the systematic and narrative reviews included in this study.

Lack of consensus on terminology and lack of precision of terms creates nosological and clinical dilemmas for researchers, clinicians and patients. In medical literature it creates difficulty identifying relevant literature. As per Castori *et al* [2] the author recommends future research use the term Generalised Joint Hypermobility (GJH) and Hypermobility Spectrum Disorders (HSD) and abandon other approximating terms.

Additionally, imprecise use of terms creates risk that hypermobility is used as a proxy term for tissue fragility, (the true marker of Ehlers Danlos Syndrome and disorders of defective collagen and connective tissue) will potentially limit diagnosis. This is an area that requires exploration in future research.

The Beighton Score has acceptable inter-rater reliability and intra-rater reliability ranges reported in the literature with the lowest ICC reported as 0.72 and 0.74 (respectively). Questions still remain regarding its validity.

Incorporation bias is a major issue of concern not previously addressed in literature to date. As such, The BS should not be used to rule out a diagnosis of GJH. In patients where there is a high level of clinical suspicion who score <4 on the BS, further clinical evaluation is warranted to ensure diagnoses are not missed as delays, or missed diagnoses can have serious negative health outcomes for patients.

There is limited high quality research available hence it is not possible to draw a conclusion about whether other tools are more or less reliable compared to BS, or valid in comparison to the BS.

No large multicentre, blinded, randomised control trial has been conducted comparing the BS to other commonly used methods, or the alternative of combination scores, or global joint ROM index.

Further research is required to quantify validity and reliability of the BS and other scoring systems. Although recommended by systematic and narrative reviews in the past, there remains paucity of literature. This should be a priority as it creates significant challenges for patients who rely on scoring systems to achieve a diagnosis of HSD/EDS and other HDCT involving GJH.

## Data Availability

All data produced in the present study are available in the supplementary materials

## REGISTRATION AND PROTOCOL

This paper was not registered as it formed part of a thesis for the University of South Wales, UK.

## SUPPORT AND CONFLICTS OF INTEREST

The author received no financial, or other type of support in undertaking this review. The author has no conflict of interest to declare.

## AVAILABILITY OF DATA AND OTHER MATERIALS

Supplementary material, and additional information can be obtained by contacting the author at the email address provided.

## LIST OF ABBREVIATIONS

GJH: Generalised Joint Hypermobility
JH: Joint hypermobility
HSD: Hypermobile Spectrum Disorder
H-EDS: Hypermobile Ehlers Danlos Syndrome
BS: Beighton Score
ROM: Range of motion
HDTC: Hereditable Disorders of Connective Tissue

## CONSENT FOR PUBLICATION

Not relevant for this review as it doesn’t contain individual data.

## FUNDING

No funding was received in the process of this review.

## CONFLICT OF INTEREST

The author has no conflict of interest to declare.

ACKNOWLEDGEMENTS

Thank you to Dr Rizwan Rajak from the University of South Wales for his supervision and support during the undertaking of this review.

## Notes

### Competing Interest Statement

The authors have declared no competing interest.

### Funding Statement

This study did not receive any funding

### Author Declarations

https://www.equator-network.org/reporting-guidelines/prisma-s/ https://link.springer.com/article/10.1007/s00296-021-04832-4

### Summary of Updates

Title modified, references edited

